# Quantitative Analysis of Dual-Task Rehabilitation

**DOI:** 10.1101/2025.03.12.25323873

**Authors:** Yutong Feng, Hongbei Meng, Zihe Zhao, Xiaomeng Wang, Xiaoxue Zhai, Yansong Hu, Guanyu Wang, Bo Peng, Wenyu Yang, Xuemeng Li, Wenxin Tao, Shuo Gao, Yu Pan

## Abstract

**BACKGROUND:** Dual-task impairment severely limits functional recovery post-stroke. This diagnostic accuracy study aimed to develop an eye-tracking-based system for objectively quantifying ankle-cognitive integration deficits in stroke survivors.

**METHODS:** This diagnostic accuracy study was conducted from January 2022 to October 2023 at Tsinghua Changgung Hospital in Beijing. A total of 20 healthy adults (mean age 53.15±6.26 years) participated in the study. In addition, 30 patients with a history of stroke (mean age 64.13±8.16 years, 8 females, disease duration 9.12±6.60 weeks) participated in a standardised dual-task evaluation. The novel system utilised 17 parameters, encompassing ankle kinematics (range of motion, velocity) and eye tracking (gaze duration, sweep latency), which were measured simultaneously during the cognitive motor task. Reliability was assessed by intragroup correlation coefficients (ICC), while criterion validity was assessed using 12 clinical evaluation metrics, including Spearman correlation with Montreal Cognitive Assessment (MOCA) scores and dual-task cost (DTC) percentages.

**RESULTS:** The system demonstrated that 88.2% of the evaluation parameters exhibited high consistency, with 55.8% showing a moderate correlation with clinical benchmark scales (p<0.05). Notably, MOCA, DTC%, and TUG-subtraction task duration were identified as key indicators of dual-task ability (P<0.05), while the Self-Rating Anxiety Scale showed lower sensitivity. Furthermore, ankle motion parameters exhibited a strong correlation with balance and fall risk (P<0.05), effectively serving as predictors of motor function recovery and fall risk in stroke patients.

**CONCLUSIONS:** This multimodal system reliably quantifies post-stroke dual-task deficits, with ankle kinematics and eye-tracking metrics serving as sensitive biomarkers for balance recovery and fall risk stratification. Findings advocate integrating objective dual-task metrics into neurorehabilitation protocols to optimize functional outcomes.

(ChiCTR2300067640; URL: https://www.chictr.org.cn/showproj.html?proj=188211).

## I. Introduction

The ability to execute dual-task or multitask execution during movement is imperative for daily life activities^1,2^. For example, walking often necessitates the maintenance of attention to changes in the surrounding environment, which facilitates appropriate gait adjustments and reduces the risk of falls. However, due to brain damage, stroke survivors, frequently experience impairments in sensory, cognitive, and motor functions, leading to a diminished capacity for dual-task execution^3,4^. Consequently, facilitating the recovery of motor-cognitive dual-task abilities in stroke patients is essential for aiding their return to normal life.

Clinical evidence suggests that motor rehabilitation training is a critical approach for restoring the functions of affected brain regions. Consequently, numerous methods incorporating motor-cognitive dual-task training have been developed to enhance dual-task performance^5^. Notably, these methods differ from traditional motor rehabilitation approaches, as dual-task training requires the design of sensory feedback-based rehabilitation paradigms to facilitate interregional brain coordination. For instance, Pelosin *et al*.^6^ introduce a treadmill training program incorporating virtual reality, in which participants are required to avoid virtual obstacles displayed on a screen while walking on a treadmill. After six weeks of training, the fall rate among Parkinson’s disease patients significantly decreases (p<0.001), along with improvements in gait speed, gait variability, and obstacle negotiation abilities. Similarly, Spanò *et al*.^7^ employ a combination of a sensory carpet and a video projector to stimulate balance, gait motor skills, and cognitive abilities. Participants are tasked with associating sounds and images, identifying numbers, and performing calculations while walking. The intervention increased from 33.8 meters in the 6-minute walking test distance and an improvement of 0.10 m/s in walking speed. Additionally, Lee *et al*.^8^ utilize robot-assisted upper-limb rehabilitation, where patients perform 640 planar reaching motions using a handle, with visual feedback provided via a display. After four weeks, patients exhibit significant enhancements in motor performance and specific cognitive test scores (p<0.01). Such dual-task-based motor paradigms effectively improve patients’ motor performance.

However, the majority of prior studies on dual-task performance have primarily focused on the upper limbs, highlighting the need for research on lower-limb motor-cognitive dual-task performance. To address this, we develop a dual-task system for ankle motor-cognitive assessment and training based on eye-tracking technology, as shown in Fig. 1. This system performs a training paradigm tailored for ankle motor-cognitive dual-tasking, where patients control a pointer on the display interface using ankle movements to sequentially select numbers, receiving real-time visual feedback through the eye-tracking interface. Furthermore, we utilize eye-tracking devices to collect realtime motion parameters of both the ankle and eye movements, that serve to reflect the patient’s ability to execute motor-cognitive dual tasks. As conventional gait parameters are insufficient cognitive effectively, for evaluating motor-dual-task performance we develop a novel assessment method by integrating existing clinical scales. To validate the clinical relevance of this method, we conduct correlation and validity analyses between the physiological parameters of eye and ankle movements and corresponding clinical benchmarks, ultimately providing an effective rehabilitation tool for stroke patients with lower-limb motor-cognitive dual-task impairments.

**Fig. 1.**
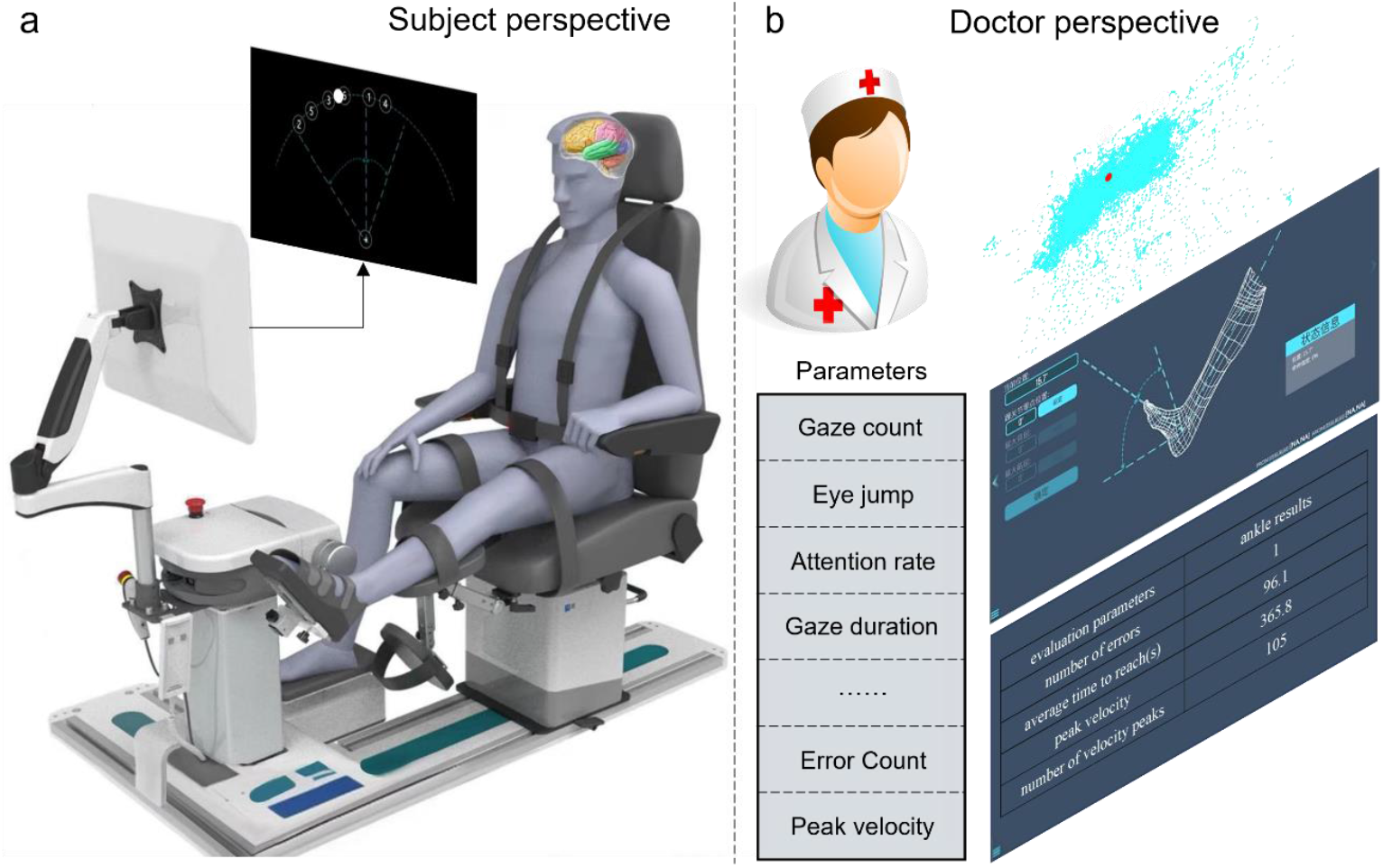
The training paradigm for dual-task ankle motor-cognitive training. (a) Subjects receive command signals for dual training of foot and ankle movement and cognition. (b) Physicians obtain eye movement information and foot and ankle movement information during training, as well as parametric indicators obtained from systematic analysis.

## II. System

### A. Training System

The system enables real-time documentation of ankle movements and eye-tracking activities of participants, providing an objective evaluation of their motor-cognitive dual-task capabilities for the ankle. The hardware foundation of this study consists of two main components: the ankle rehabilitation robot and an eye-tracking device equipped with a display screen.

The ankle rehabilitation robot is an anti-spasticity stretching device, capable of achieving a maximum dorsiflexion angle of 25° and a maximum plantarflexion angle of 45°. It employs a dual “mechanical + software” limiting mechanism to ensure operational safety.

The eye-tracking device, named the 4L Eye Tracker, utilizes a high-precision eye movement detection system powered by the Tobii Stream Engine API to generate eye-tracking data streams. The corneal reflection eye tracker is positioned 60–70 cm in front of the patient to ensure accurate and reliable data collection. Key features include: 1) support for dual-screen monitoring; 2) a resolution of 1920 × 1080; 3) an auto-start eye-tracking service (port 9000, UDP protocol, WiFi-based connection); 4) a defined area of interest corresponding to the optimal tracking range, where the gaze region falls within 56%–64% of the screen; and 5) the ability to track eye and head (gaze) movements over a long distance with a maximum sampling frequency of 60 Hz and an accuracy of 0.5 degrees. Additional technical specifications are detailed in Table I.

**TABLE I.**
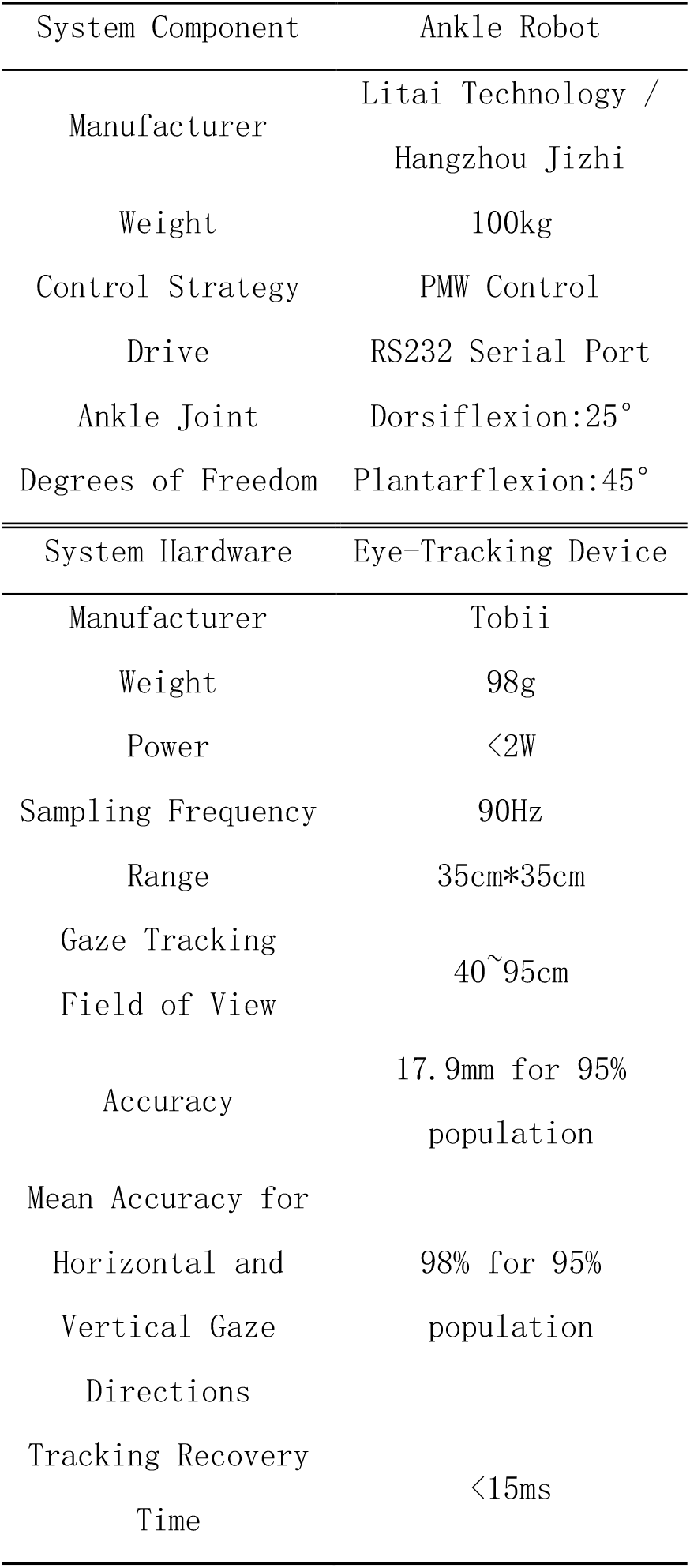
Hardware Information.

The ankle rehabilitation robot is equipped with position sensors to collect parameter data during ankle movements.

While eye-tracking device incorporates visual sensors to capture eye movement data. The processing unit analyzes the participant’s attentional state during ankle joint movements based on eye-tracking data, enabling the synchronous collection of both ankle motion and eye movement data during task execution. A non-contact connection is employed between these two components, ensuring that the stability of the eye-tracking device remains unaffected when the ankle robot facilitates joint movements.

### B. Testing Procedure

The participant is seated 60–70cm away from the eye-tracking screen, with the height and backrest distance of the specialized experimental chair adjusted according to their comfort. The tested lower limb is positioned with the knee joint fully extended, while the ankle joint is placed in the neutral position on the foot support of the ankle robot and securely fixed with straps.

Eye-tracking calibration is performed before each evaluation test. During the calibration process, a total of seven points are presented. The participant is required to follow the prompts and fixate on each point until it disappears, completing the calibration process.

Subsequent to the initiation of the software, the device automatically performs a mechanical limit detection. The operator must set the ankle joint’s neutral position based on the participant’s range of motion and measure the maximum active dorsiflexion and plantarflexion motion angle.

After commencing the Trail Making Test (TMT), the participant is first familiarised with the target paradigm through a pretest. The participant is instructed to perform foot movements in numerical order, guiding a white circle to align with the corresponding numbered circle, as shown in Fig. 2. A green indicator signifies success, while a red indicator signifies failure, requiring the participant to return to the red area and continue the test. Following the pretest, the formal test is conducted with a 2-minute interval between tests on each side. As illustrated in Fig. 3, the overall training process is comprised of a series of distinct stages.

**Fig. 2.**
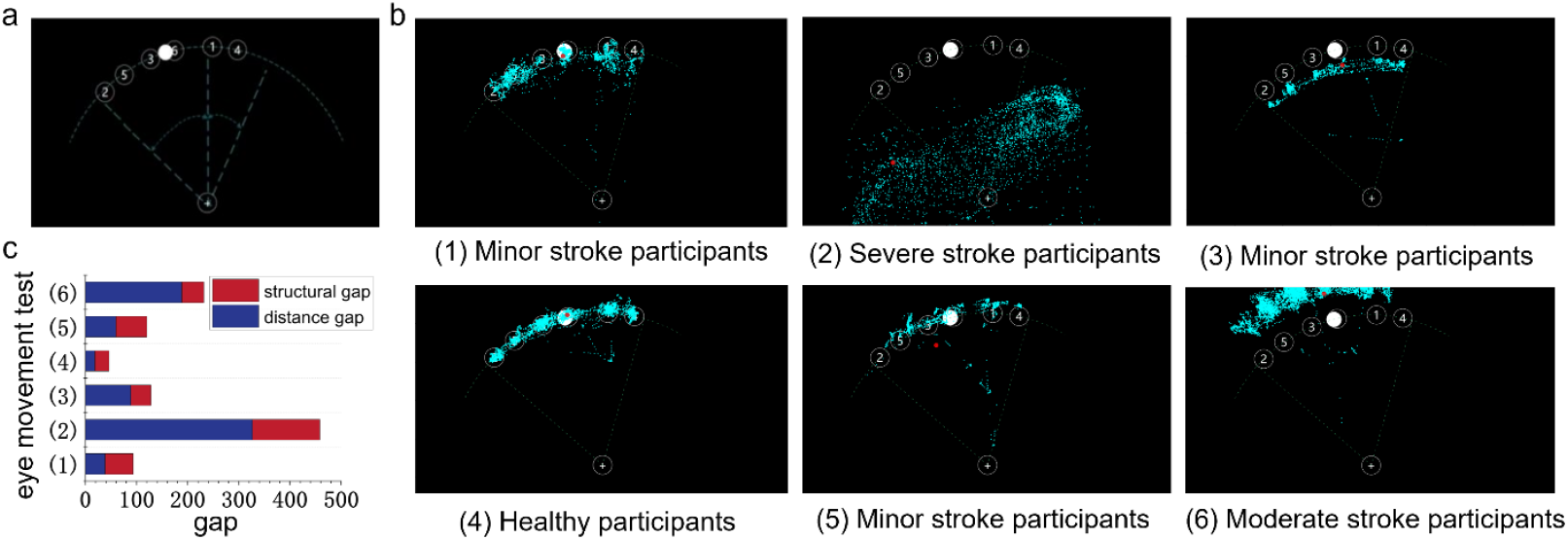
Eye movement capture during motor cognition training. (a) The motor cognition paradigm of the TMT mission test, in which subjects were required to move sequentially along an arc trajectory starting from the white dot to the specified numbered dots. (b) Six typical subjects’ eye movements capture dots. (c) Histograms of structural and distance differences in the set of eye movement dots versus the standard arc for these six subjects.

**Fig. 3.**
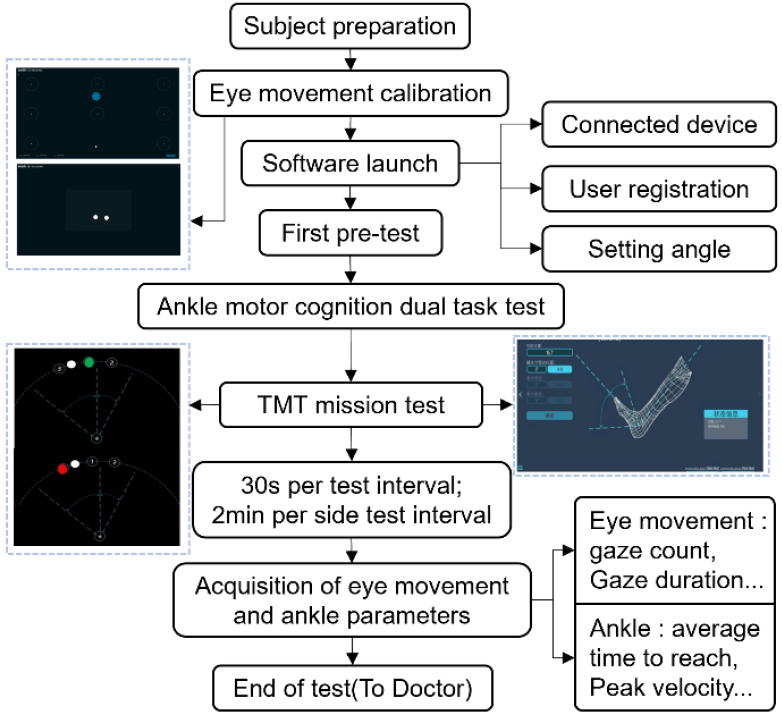
Training Paradigm System Process for TMT mission test.

## III. Method

This study recruited 20 healthy adult participants from Beijing Tsinghua Changgung Hospital between January 2022 and October 2023. The dominant foot of each healthy participant was subjected to the ankle motor-cognitive dual-task test, followed by a retest one week later. The reliability of the test was evaluated through a reliability study by analyzing the intraclass corre-lation coefficient (ICC) between the two test results. Additionally, 30 inpatients diagnosed with stroke in the Department of Rehabilitation Medicine at Beijing Tsinghua Chang-gung Hospital during the same period were recruited. These stroke patients underwent ankle motor-cognitive dual-task testing on both sides. The validity of the test was assessed through a validity study by analyzing the correlation between the test results and clinical functional benchmarks.

### A. Inclusion and Exclusion Criteria

#### 1) Healthy Participants

##### Inclusion Criteria

a. Individuals with a history of good physical health;
b. Unrestricted ankle joint mobility;
c. Normal binocular visual field;
d. Age between 30 and 75 years.

##### Exclusion Criteria

a. Individuals with cognitive, speech, or mental disorders that prevent cooperation;
b. Lower limb fractures or similar conditions;
c. Recent use of stimulants or depressants affecting the nervous system;
d. Inability to complete the entire evaluation and testing process.

#### 2) Stroke Patients

##### Inclusion Criteria

a. First-time onset of stroke, with lesions confined to the middle cerebral artery or vertebrobasilar artery system; stroke types include cerebral infarction, cerebral hemorrhage, and cerebral embolism.
b. Stroke onset between 1 and 6 months before the study.
c. Manual Muscle Test (MMT) score > 3 for the affected lower limb (including ankel plantarflexion and dorsiflexion).
d. Brunnstrom stage ≥ 4 for the affected lower limb, with at least one item scoring < 5 on the Stroke Impact Scale (SIS) for the lower limb.
e. Normal binocular visual field.
f. Age between 30 and 75 years.

##### Exclusion Criteria

a. Co-occurrence of other neurological diseases.
b. Severe cognitive impairments, including a Montreal Cognitive Assessment (MOCA) score < 20, significant hemispatial neglect, body agnosia, or apraxia.
c. Severe aphasia, mental disorders, or emotional disturbances that prevent cooperation.
d. Lower limb fractures or similar conditions.
e. Poor compliance, as judged by the researchers, makes it impossible to complete the study as required.

##### Discontinuation Criteria

a. The participant experiences a fall during the trial (a fall is defined as an unintended descent to the ground, floor, or another lower level)^9^.
b. The participant is unable to complete the trial continuously in one session.
c. The participant withdraws from the trial midway.

This study was reviewed and approved by the Ethics Committee of Beijing Tsinghua Changgung Hospital (Approval Number: 22207-0-02), and informed consent was obtained from all participants.

### B. Pilot Study

The sample size estimation for this study was performed using PASS 15.0 software with a significance level of α=0.05. A single-sample mean approach was used for the calculation, based on the primary parameter “fixation count (FC)” obtained from the ankle motor-cognitive dual-task test. A preliminary experiment was conducted with three healthy participants, yielding an initial standard deviation of 1.14 for FC. With an allowable error of 0.5, the final calculation indicated that at least 20 healthy participants were required. Similarly, a preli-minary experiment involving three stroke patients resulted in an initial standard deviation of 1.02 for FC. Using the same allowable error of 0.5, the final calculation determined that at least 30 stroke patients were needed.

### C. Observation Indicators

#### 1) Eye-Tracking Parameters

Primary Indicators: FC, saccade amplitude (SA).

Secondary Indicators: First fixation duration (FFD), percentage of saccades before peak velocity (S-BPV%), percentage of saccades after peak velocity (S-APV%), fixation rate (FR), gaze duration (GD), the spatial density of fixations (SDFs), number of saccades reaching the target (NS-RT), the average number of stationary saccades (ANSS).

#### 2) Ankle Movement Parameters

Primary Indicators: Total range (TR), average speed (AS).

Secondary Indicators: Total duration (TD), error count (EC), the average time to target (ATT), peak velocity (PV), and number of velocity peaks (NVPs).

#### 3) Clinical Evaluation Indicators

Primary Indicators: MOCA score, Mini-Mental State Examination (MMSE) score, Fugl-Meyer Assessment - Lower Extremity (FMA-LE) score, 10-meter walking average speed (10mAS), Timed Up and Go Test (TUG) / TUG-subtraction task duration (TUG-STD), dual-task cost percentage (DTC%). Secondary Indicators: Berg Balance Scale (BBS) score, Morse Fall Risk Assessment (MFRA) score, Holden Walking Ability (HWA) scale, Modified Barthel Index (MBI), Self-Rating Anxiety Scale (SAS) score, Self-Rating Depression Scale (SDS) score.

## IV. Results

### A. Test-Retest Reliability

The test-retest reliability of the dominantside ankle TMT task among healthy participants revealed good to excellent consistency^10^ for 10 eye-tracking parameters(FC, FFD, GD, FR, SDFs, NS-RT, ANSS, SA, ATT) ranged from 0.601 to 0.897, and moderate to good consistency for 5 ankel plantar and dorsal extension movement parameters (TR, AS, EC, NVPs, PV, S-BPV%, and S-APV%) ranged from 0.509 to 0.639. However, EC and TD parameters exhibited lower consistency across the two tests, with ICC values ranging from 0.319 to 0.375, as shown in Fig. 4(a).

**Fig. 4.**
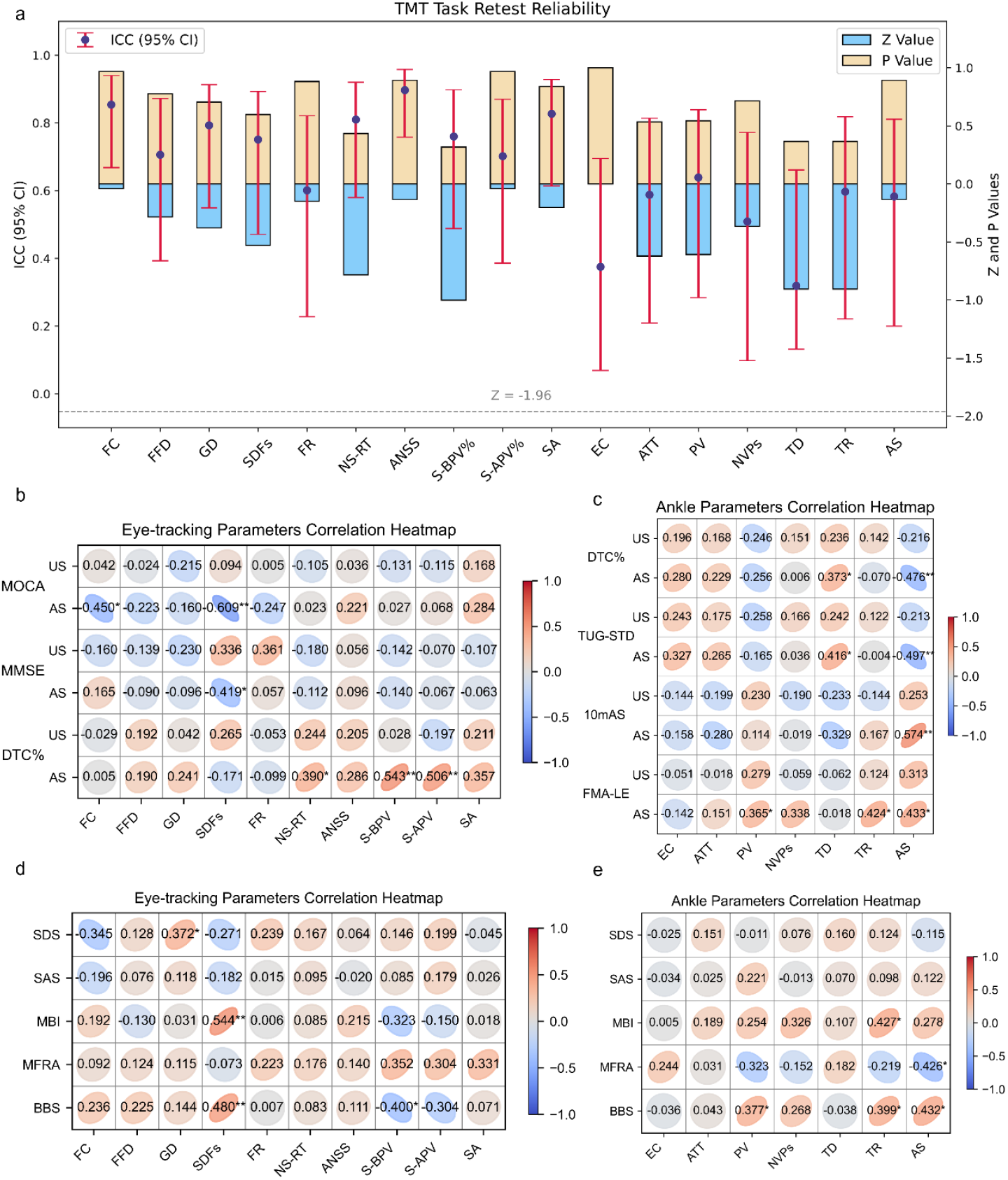
Retest reliability and correlation analysis of motor-cognition dual-task parameters in post-stroke individuals. (a) Retest reliability of TMT task parameters with ICC (95% CI) (interval plots), Z-scores (blue bars), and P-values (yellow bars). (b) Correlation heatmap of eye-tracking parameters with MOCA, MMSE, and DTC scores for US and AS sides. (c) Correlation heatmap of ankle parameters with FMA-LE, 10mAS, TUG-SC, and DTC scores for US and AS. (d)Correlation heatmap of eye-tracking parameters with BBS, MFRA, TMT, SAS, and SDS scores. (e)Correlation heatmap of ankle parameters with BBS, MFRA, TMT, SAS, and SDS scores.

### B. Concurrent Criterion Validi

This test aimed to evaluate the correlation between test results and clinical indicators. Therefore, specific clinical func-tional benchmarks were selected for validating the system’s effectiveness for both eye-tracking and ankle parameters. For eye-tracking parameters, the associated benchmarks included the MMSE score, MOCA score, and DTC%. For ankle parameters, the related benchmarks included the FMA-LE score, 10mAS, TUG-STD, and DTC%, as detailed in Table II.

**TABLE II.**
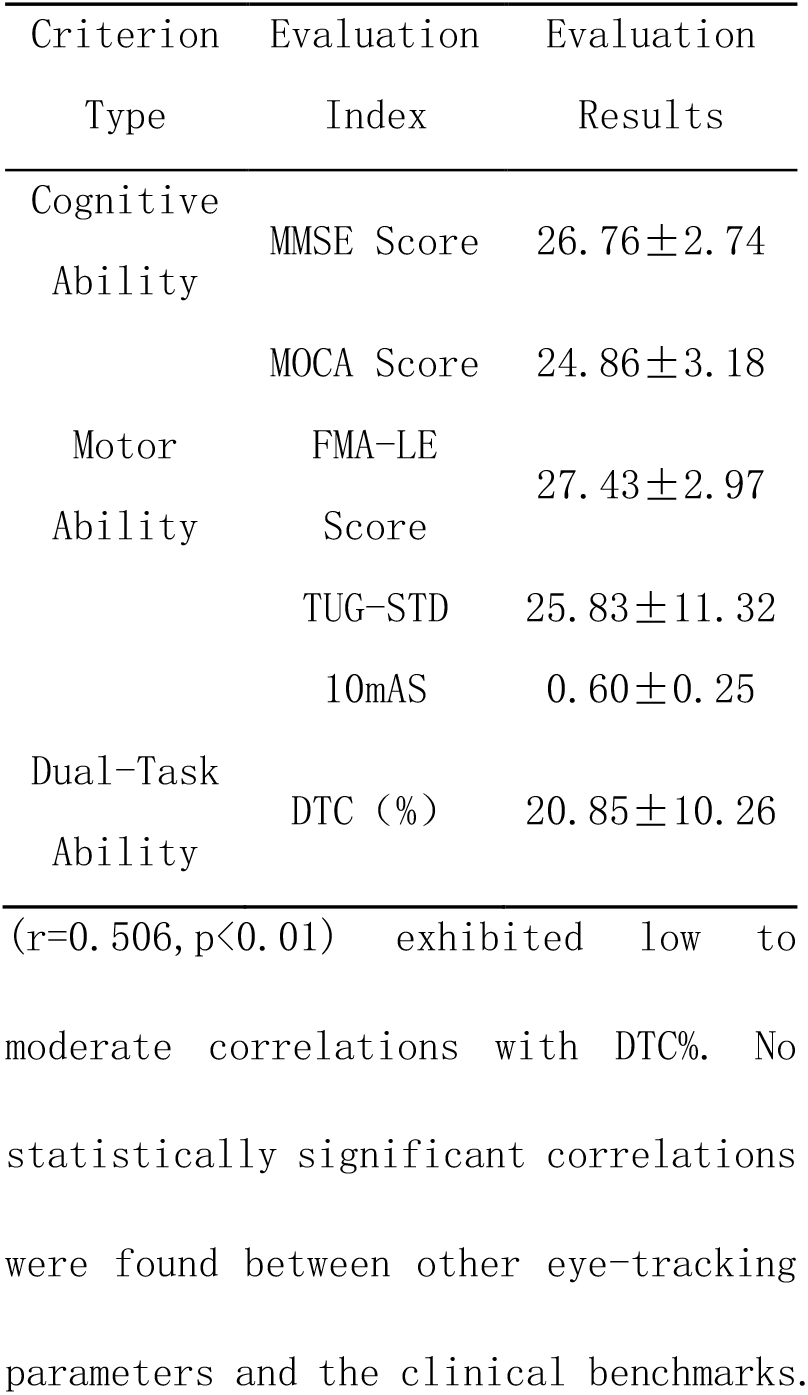
Criterion Evaluation Results of Stroke Patients.

The correlation analysis of eye tracking parameters is shown in Fig. 4(b). A moderate correlation between FC and MOCA score (r=-0.450, p<0.05). SDFs showed moderate correlations with both MMSE score (r=-0.419, p<0.05) and MOCA score (r=-0.609, p<0.01). Additionally, NS-RT (r=0.390, p<0.05), S-BPV% (r=0.543, p<0.01), and S-APV% (r=0.506,p<0.01) exhibited low to moderate correlations with DTC%. No statistically significant correlations were found between other eye-tracking parameters and the clinical benchmarks.

The correlation analysis of ankle movement parameters is shown in Fig. 4(c). PV (r=0.365, p<0.05) and TR (r=0.424, p<0.05) show low to moderate correlations with FMA-LE scores, as does TD with TUG-STD (r=0.416, p<0.05) and DTC% (r=0.373, p<0.05). The correlation of AS with FMA-LE scores (r=0.433, p<0.01), 10mAS (r=0.574, p<0.01), TUG-STD (r=-0.497, p<0.01) and DTC% (r=-0.476, p<0.01) showed a moderate correlation.

#### 2) Performance Results for the Unaffected Side

For the unaffected side of stroke patients during the TMT task, no statistically significant correlations were observed between the eye-tracking parameters and their three benchmarks, nor between the ankle parameters and their four benchmarks. Some of the resulting parameters are shown in Fig. 5.

**Fig. 5.**
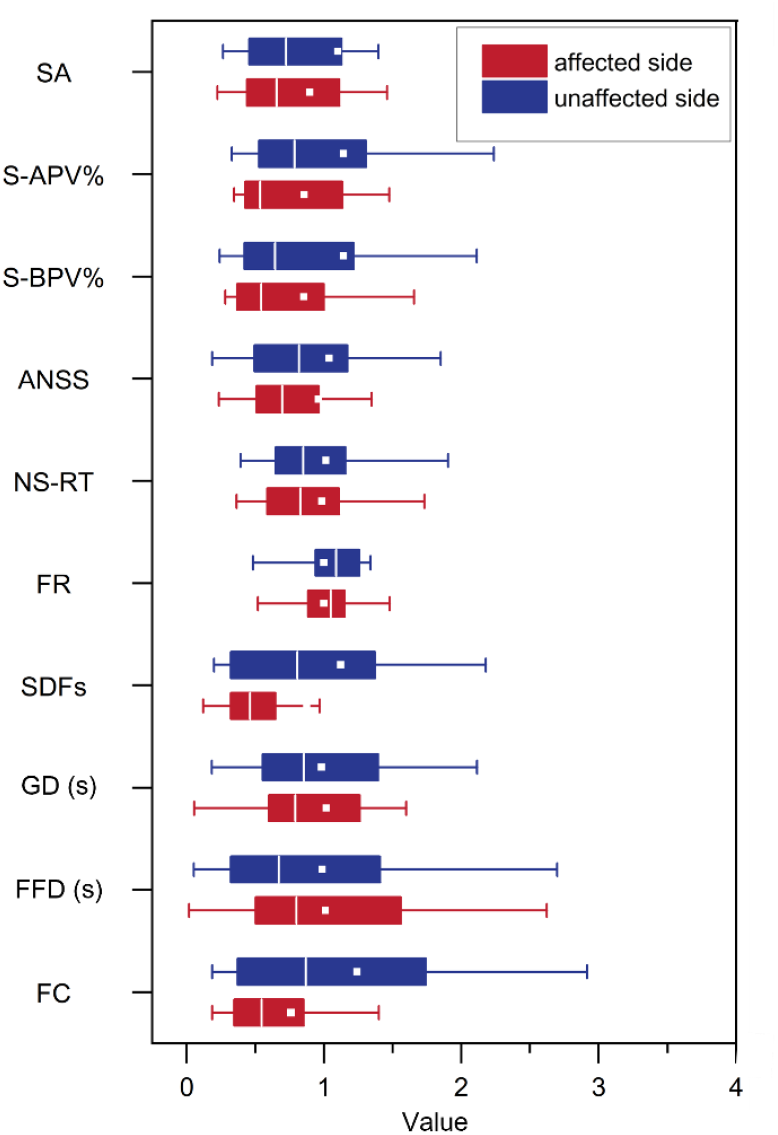
Box plot of the averaged results of some parameter indicators in the dual-task training of ankle movement and cognition in the unaffected side (US) and the affected side (AS) of stroke patients.

### C. Correlation with Clinical Scale Evaluations

Clinical functional evaluations primarily included assessments of balance ability (BBS score, MFRA score), activities of daily living (MBI), and emotional state (SAS score, SDS score), as shown in Table III. The correlation between the evaluation parameters and clinical functional assessment is shown in Fig. 4(d)(e).

**TABLE III.**
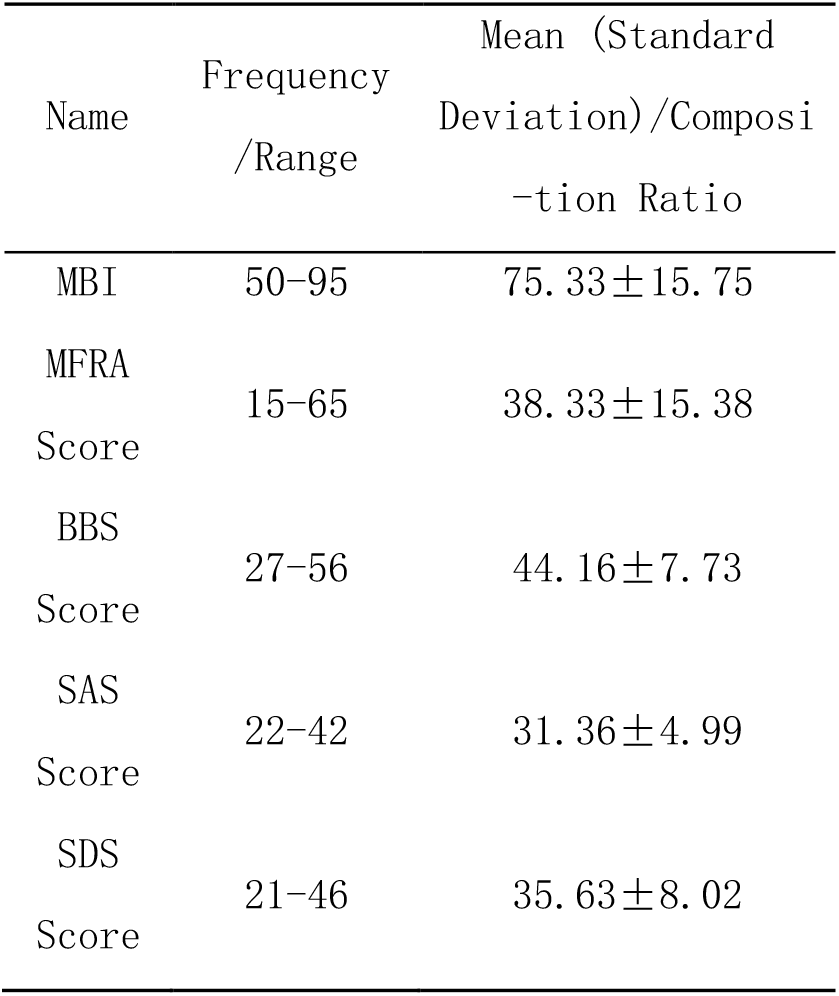
Clinical Scale Evaluation Results of Stroke Patients (N=30)

#### 1) Balance Ability

Eye-tracking parameters revealed a moderate positive correlation between the SDFs and BBS scores (r=0.480, P<0.01), indicating that higher SDFs correspond to better balance ability. Conversely, S-BPV% showed a moderate negative corre-lation with BBS scores (r=-0.400, P < 0.01), suggesting that a higher S-BPV% is associated with poorer balance ability.

Ankle assessment parameters showed a low positive correlation between PV and BBS scores (r=0.377, p<0.05), indicating that higher PV corresponds to better balance ability. Similarly, TR exhibited a low positive correlation with BBS scores (r=0.399, p<0.05), suggesting that greater TR is associated with improved balance ability. AS demonstrated a moderate positive correlation with BBS scores (r=0.432, p<0.05), indicating that faster AS is linked to better balance ability. Additionally, AS showed a moderate negative correlation with MFRA scores (r=-0.426, p<0.05), meaning that faster AS is associated with a lower risk of falls.

#### 2) Independence in Activities of Daily Living

Eye-tracking parameters showed a moderate positive correlation between the SDFs and MBI (r=0.544, p<0.01), indicating that higher SDFs are associated with greater independence in activities of daily living.

Ankle assessment parameters demonstrated a moderate positive correlation between TR and MBI (r=0.427, p<0.05), indicating that a greater TR is associated with higher independence in activities of daily living.

#### 3) Emotion

Eye-tracking parameters showed a low positive correlation between GD and SDS scores (r=0.372, p<0.05), indicating that longer GD is associated with more severe depressive symptoms.

Ankle assessment parameters from the TMT task showed no significant correlations with emotional state indicators.

## V. Discussion

### A. Test-Retest Reliability

This study assessed the reliability of the ankle motor-cognitive dual-task system, confirming that most parameters demonstrate robust test-retest reliability, with ICC values ranging from 0.509 to 0.897. However, the parameter TD, which represents the total time required to complete the dual-task paradigm, showed poor test-retest reliability (ICC < 0.40). This may be due to the lack of time constraints or defined time intervals during testing, as well as the influence of confounding factors, which could reduce its stability in reflecting participants’ processing or execution abilities. To enhance its reliability, future studies should better control variables such as test duration, environmental conditions, and participant states to ensure more robust validation of this parameter in ankle motor-cognitive dual-task assessments.

Eye-tracking technology, leveraging its unique strengths, offers profound insights into the nuanced characteristics of cognitive processing and cognitive load through meticulous analysis of eye-tracking data^11^. J. M. Wolfe *et al*.^12^ revealed that cognitive models governing visual attention during visual search tasks are fundamentally based on fine-grained analyses of visual features.

This discovery not only advances our understanding of visual attention mechanisms but also establishes a solid theoretical foundation for applying eye-tracking technology in cognitive science research. The TMT task paradigm, as a classical visual search task, employs indicators such as FC, FFD, FR, SDFs, SA, GD, NS-RT, number of stationary saccades, S-BPV%, and S-APV%. These metrics primarily reflect cognitive features such as search efficiency, attention allocation, processing capacity, and cognitive load. For example, FC indicates the number of times the eyes remain fixed on a target, whereas higher FC suggests greater difficulty in identifying the target, lower search efficiency, and higher cognitive load^13^. In the test-retest reliability analysis of the ankle motor-cognitive dual-task test, these eye-tracking parameters demonstrated strong intraclass consistency, indicating their stability in reflecting participants’ cognitive processing characteristics. Similarly, ankle motion parameters, including EC, ATT, PV, NVPs, TR, and AS, primarily reflect processing and execution capabilities. For instance, AS is calculated as the angle or distance moved by the ankle from the starting position to the endpoint divided by the total time taken, where higher AS indicates better information processing and execution capabilities. The study results demonstrated that these ankle motion parameters also exhibited good test-retest reliability, confirming their stability in reflecting participants’ motor characteristics during the execution of ankle motor-cognitive dual-task tests.

Test-retest reliability is also influenced by the retest interval. Studies on dual-task or robotic quantitative assessments have employed retest intervals ranging from 1 hour^14^, 2 hours^15^, 1 week^16,17^, to 2 weeks^18^. Longer intervals increase measurement variability and reduce test-retest reliability, while shorter intervals may introduce fatigue and learning effects, impacting the results. Therefore, selecting an appropriate retest interval is crucial for the analysis of test-retest reliability. Lauren Rachal *et al*.^17^ investigated the reliability and clinical feasibility of measuring dual-task gait in inpatient rehabilitation settings for patients with traumatic brain injury, using a 1-week retest interval, which demonstrated excellent reliability. Based on prior studies and considering fatigue and memory effects after testing, this study also adopted a 1-week retest interval, resulting in good test-retest reliability for most parameters. Additionally, the proactive nature of the ankle motor-cognitive dual-task test (providing pretest guidance to participants) offers an advantage over previous dual-task tests by eliminating the need to withhold test details from participants, thereby enhancing repeatability.

### B. Concurrent Criterion Validity

Concurrent criterion validity is used to determine whether the ankle motor-cognitive dual-task test for stroke patients reflects their motor and cognitive conditions in a manner consistent with clinical benchmark assessments.

The MMSE and MOCA are widely used cognitive screening tools in clinical practice^19^, covering domains such as orientation, memory, attention, calculation, and visuospatial skills. Both tools are primarily used for preliminary screening. Recent years have seen dual-task assessments demonstrate significant advantages in identifying cognitive deficits in highfunctioning stroke patients, increasingly applied in clinical evaluation and cognitive training. Songjun Lin *et al*.^20^ showed that dual-task assessments based on the Stroop paradigm more effectively identify cognitive deficits in high-functioning stroke patients, particularly in attention evaluation, with greater sensitivity and specificity.

Our study found that the FC was moderately correlated with MOCA scores, while the spatial density of fixations was moderately correlated with both MMSE scores and MOCA scores. These demonstrate that eye-tracking parameters were consistent with MMSE and MOCA evaluation results and reflect the cognitive functioning level of patients when performing dual-tasks.

The FMA scale is a comprehensive tool for evaluating motor function in stroke patients^21,22^, particularly suitable for those with moderate motor function levels. It includes 17 lower-limb assessment items that are closely related to activities of daily living, although it may be influenced by the subjective judgment of the evaluator. 10mAS and TUG-STD are direct indicators of lower-limb motor ability. Studies have shown that AS has a moderate correlation with these benchmarks, indicating that greater AS reflects stronger lower-limb motor function. The unique characteristics of ankle parameters under dual-task conditions and the singularity of these benchmarks may explain the lower correlation between other ankle parameters and the benchmarks.

Moreover, DTC% quantifies the degree of dual-task interference by calculating the task effects for each task. This metric is commonly used in research to evaluate dual-task performance^23^. In this study, it was calculated using the results of the TUG test and TUG-STD^24^. Research has shown that lower cognitive ability is often associated with higher DTC%^25^, indicating that individuals with relatively poor cognitive function may face greater challenges when managing two tasks simultaneously. This finding aligns with the results of this study and provides valuable insights into the mechanisms of dual-task motor-cognitive interactions. It also offers critical guidance for developing personalized training programs tailored to individuals with varying levels of motor-cognitive dual-task capabilities.

Overall, compared to the unaffected side, the parameters from the affected side demonstrated strong-er correlations with clinical benchmarks, likely due to the asymmetry in kinematic characteristics observed after stroke. The unaffected side of stroke patients experiences relatively less impairment than the affected side, resulting in weaker correlations with benchmark outcomes. As noted by Jeong-Woo Seo *et al*.^26^, hemiparetic stroke patients exhibit asymmetrical kinematic changes, characterized by distinctions between the unaffected and affected sides, which can be quantified using symmetry ratios. Additionally, the lack of significant validity for parameters from the unaffected side, compared to the significant validity of parameters from the affected side, indirectly highlights the differences between the two sides. Currently, quantitative indicators capable of reflecting the bilateral differences in stroke patients are insufficient. Therefore, further analysis of the ankle motor-cognitive dual-task parameter characteristics for each side in stroke pati-ents is needed to identify sensitive parameters that can effectively detect or differentiate bilateral asymmetries, paving the way for the development of a novel evaluation method.

In summary, the validity study clarified the effectiveness and practical value of the evaluation parameters in the test. FC and SDFs effectively reflect the cognitive abilities of stroke patients, revealing specific eye-tracking characteristics in dual-task scenarios and uncovering internal cognitive load. Similarly, AS effectively reflects lower-limb motor ability and dual-task cost, highlighting the ankle parameter characteristics in dual-task scenarios, revealing internal dual-task inter-ference effects and motor insufficien-cies, and providing guidance for the rehabilitation treatment of clinical stroke patients. Furthermore, the results of the criterion validity study offer references for the selection of benchmark tools in future dual-task validity studies based on eye-tracking technology. The innova-tive task design, comprehensive test content, and objective results of the motor-cognitive dual-task test based on eye-tracking technology demonstrate its sensitivity, quantifiability, ecological validity, and dual-task performance insights. In contrast, traditional clinical evaluation scales are limited by ceiling effects, simplistic test content, subjective results, and poor sensitivity to mild functional impairments. Therefore, when conducting criterion validity studies, highly consistent benchmark tools, such as dual-task gait tests, should be selected to optimize validity evaluation outcomes.

### C. Correlation Analysis

#### 1) Correlation with Balance and Fall Risk

Cognition and motor control share common brain networks, particularly in the prefrontal and temporal regions involved in performing cognitive and motor control tasks simultaneously under the limited capacity model, which can lead to network overload. This makes dual-task testing an effective method for the early detection of abnormalities in brain functional networks^27^ and a predictor of fall risk in stroke patients^28^. Research has shown a strong relationship between visual search and reaching movements following a stroke^29^, with stroke patients performing TMT tasks exhibiting frequent saccades (rapid eye movements), and these excessive saccades indicating a decline in dual-task performance^30^. BBS, commonly used to assess dynamic balance and predict fall risk^31^, shows that higher BBS scores correlate with better balance ability and lower fall risk^32,33^. In the TMT task, AS was identified as the ankle parameter most strongly correlated with both BBS and MFRA scores, where greater AS in the ankle motor-cognitive dual-task test was associated with higher BBS scores, lower MFRA scores, improved balance, and reduced fall risk. Additionally, balance and walking speed are critical for functional mobility, including gait and transfers, with the BBS and walking speed frequently used clinically to evaluate mobility^34,35^. Impaired balance control and slower walking speed negatively affect functional mobility, emphasizing the importance of these parameters in stroke rehabilitation.

#### 2) Correlation with Independence in Activities of Daily Living

MBI is commonly used in clinical practice to measure activities of daily living (ADL)^36^, encompassing tasks such as eating, bathing, dressing, personal hygiene, bowel and bladder control, toileting, transferring, and walking. Each activity is rated on a five-level scale, with different levels representing varying degrees of independence; higher levels indicate greater independence. A higher total MBI score reflects better ADL capability and less severe functional impairment. Accordingly, parameters such as TR and SDFs can provide insights into a patient’s level of independence in daily activities and the extent of functional disability, offering valuable guidance for clinical rehabilitation assessments and the formu-lation of rehabilitation plans.

#### 3) Correlation with Emotion

Studies have shown that approximately one-third of stroke patients experience depression, which significantly hinders functional recovery, reduces quality of life, and is closely associated with high mortality rates^37^. Emotion, as a complex psychological state, encompasses multiple components, including subjective experience, psychophysiological responses, behavioral expressions, and cognitive processes^38^, with five key elements: cognitive appraisal, physical symptoms, action tendencies, expression, and feelings^39^. Ling Luo *et al*.^40^ found that emotional states are related to cognitive impairments. In recent years, some studies have used eye-tracking data features, such as fixation duration and saccades, to identify emotional states^41^. Additionally, research by Kyoungah Kim *et al*.^42^ demonstrated that motor-cognitive dual-task training programs significantly reduce depressive symptoms. However, effective features of eye movement for emotion recognition have been explored in only a limited number of studies, and these features remain difficult to quantify or standardize. The results of this study revealed varying degrees of correlation between parameters from different ankle motor-cognitive dual-task tests and emotional scale scores in stroke patients, providing valuable references for identifying emotion-related eye-tracking features. However, the paradigm used in this study was a classical visual search task, resulting in dispersed eye-tracking parameters for emotion, making them challenging to capture accurately. In the future, we aim to refine our emotion research paradigm to extract more meaningful eye-tracking features and conduct emotion classification studies, enabling a clearer understanding of the effec-tiveness of various eye-tracking features in emotion recognition tasks. This will deepen our understanding of emotional changes in stroke patients and provide stronger support for clinical evaluation and treatment.

In summary, the results of this study indicate that different parameters from various tests show varying degrees of positive or negative correlations with clinical scale evaluations in stroke patients, preliminarily validating the clinical utility of these parameters. However, due to the richness of the test content and the large number of evaluation parameters obtained, the current guidance for clinical functions remains somewhat fragmented, and the relative weights of the parameters are not yet well-defined. Future studies could expand the sample size and employ computational approaches such as deep learning to classify, weigh, and integrate the evaluation results of stroke patients, ultimately establishing a comprehensive assessment model for predicting patient functional levels.

## VI. Conclusion

We develop a system for ankle joint movement cognition, utilizing dual-task technology based on eye-tracking technology. The reliability and validity of the system are analyzed, and correlation studies are conducted with clinical scale evaluation results. The results demonstrate that 88.2% of the evaluation parameters exhibited good test-retest consistency, and the majority of indicators exhibit moderate or above correlation with the clinical scale. Additionally, the analysis indicates that scales such as BBS can effectively reflect dual-task execution ability and can be classified as good evaluation criteria. The system’s clinical applications include its use in guiding rehabilitation plans and evaluating outcomes for stroke patients, thus providing a novel approach to balance ability and fall risk prediction.

## Data Availability

Funder requirements are not applicable, and data will not be shared or will be available upon request.

## Nonstandard Abbreviations and Acronyms

ICC: intragroup correlation coefficients
MOCA: Montreal Cognitive Assessment
DTC: dual-task cost
TMT: Trail Making Test
SIS: Stroke Impact Scale
FC: fixation count
SA: saccade amplitude
FFD: first fixation duration
S-BPV%: saccades before peak velocity
S-APV%: saccades after peak velocity
FR: fixation rate
GD: gaze duration
SDFs: the spatial density of fixations
NS-RT: number of saccades reaching the target
ANSS: average number of stationary saccades
TR: total range
AS: average speed
TD: total duration
EC: error count
ATT: the average time to target
PV: peak velocity
NVPs: number of velocity peaks
MMSE: Mini-Mental State Examination
FMA-LE: Fugl-Meyer Assessment – Lower Extremity
10mAS: 10-meter walking average speed
US: unaffected side
TUG: Timed Up and Go Test
STD: subtraction task duration
BBS: Berg Balance Scale
MFRA: Morse Fall Risk Assessment
HWA: Holden Walking Ability
MBI: Modified Barthel Index
SAS: Self-Rating Anxiety Scale
SDS: Self-Rating Depression Scale
AS: affected side
ADL: activities of daily living
MMT: Manual Muscle Test

